# The French Covid-19 vaccination policy did not solve vaccination inequities: a nationwide longitudinal study on 64.5 million individuals

**DOI:** 10.1101/2022.01.03.22268676

**Authors:** F. Débarre, E. Lecoeur, L. Guimier, M. Jauffret-Roustide, A.-S. Jannot

## Abstract

**Context:** To encourage Covid-19 vaccination, France introduced during the Summer 2021 a “Sanitary Pass,” which morphed into a “Vaccine Passe” in early 2022. While the Sanity Pass led to an increase in Covid-19 vaccination rates, spatial heterogeneities in vaccination rates remained. To identify potential determinants of these heterogeneities and evaluate the French Sanitary and Vaccine Pass’ efficacies in reducing them, we used a data-driven approach on exhaustive nationwide data, gathering 141 socio-economic, political and geographic indicators.

**Methods:** We considered the association between being a district above the median value of the first-dose vaccination rates and being above the median value of each indicator at different time points: just before the sanitary pass announcement (week 2021-W27), just before the sanitary pass came into force (week 2021-W31) and one month after (week 2021-W35), and the equivalent dates for the vaccine pass (weeks 2021-W49, 2022-W03, 2022-W07). We then considered the change over time of vaccination rates according to deciles of the three of the most associated indicators.

**Results:** The indicators most associated with vaccination rates were the share of local income coming from unemployment benefits, the proportion of overcrowded households, the proportion of immigrants in the district, and vote for an “anti-establishment” candidate at the 2017 Presidential election. Vaccination rate also were also contrasted along a North-West – South East axis, with lower vaccination coverage in the South-East of France.

**Conclusion:** Our analysis reveals that, both before and after the introduction of the French sanitary and vaccination passes, factors with the largest impact are related to poverty, immigration, and trust in the government.

## Introduction

The rapid development of effective COVID-19 vaccines brought the hope of a rapid return to normalcy, but heterogeneous vaccination rates, both among countries because of inequitable distributions of doses (Usher 2021) and within countries (Caspi et al. 2021; Murthy et al. 2021), jeopardize epidemic control.

Hesitancy and hostility toward vaccination have been comparatively high in France in recent decades (European Commission. Directorate General for Health and Food Safety. 2018). Modern vaccine hesitancy in France started with claims of a link between the hepatitis B vaccine and multiple sclerosis (J. K. Ward et al. 2019); it strongly increased following the 2009-2010 vaccination campaign against pandemic flu, the contested management of which in France was a tipping point that led to higher vaccine hesitancy and hostility (Guimier 2021; J. K. Ward et al. 2019). The trend was confirmed with the COVID-19 pandemic (Lindholt et al. 2021; Spire, Bajos, and Silberzan 2021): just before Covid-19 vaccines became available, intentions to get vaccinated were comparatively very low in France compared to other countries (44% of the respondents in (Wouters et al. 2021) in the Fall 2020; about 40% of respondent in (Santé Publique France 2021) in December 2020). Acceptance of the COVID-19 vaccine however gradually grew during 2021 (Santé Publique France 2021; J. Ward 2021).

Spatial heterogeneties in vaccination rates have already been documented in France for previous vaccines. Vaccination coverage for the Hepatitis B vaccine and for the Measles-Mumps-Rubella vaccine has been lower in the South of France, and especially in the SouthEast of the country (Guimier 2021). Distance to the central political power in Paris, as well as a sense of belonging to a local community with a strong cultural identity, have been put forward as potential explanations for this geographic gradient in vaccination rates (Guimier 2021).

Attitudes toward vaccination are also known to be influenced by social and territorial inequalities. Surveys conducted in 2020 in France showed that respondents with lower education (Schwarzinger et al. 2021; Coulaud et al. 2022), lower income levels or less trust in authorities (Spire, Bajos, and Silberzan 2021; Lindholt et al. 2021) were more likely to be hostile to COVID-19 vaccines.

By mid-July 2021, France was facing an epidemic wave due to the Delta variant. To speed up vaccination, President Macron announced on 12 July 2021 the implementation of a domestic “sanitary pass” (le passe sanitaire), which came fully into force on 9 August 2021. Presenting as a QR code, a long-term sanitary pass was obtained after full vaccination (two doses, or only one dose in the case of a documented previous Covid-19 infection), and a short-term version could be obtained with a negative Covid-19 test. The “sanitary pass” was required in most cultural venues, for both indoor and outdoor dining and in health structures. This announcement led to an unprecedented demand for vaccination (Oliu-Barton et al. 2022), which was considered internationally as a potential model to follow. Vaccination rates climbed from about 64% of the population over 20 years old by 11 July 2021 (52% of all ages) to 82 on 5 September 2021 (69% of all ages). Because it targeted pay-for social activities, however, the “sanitary pass” was feared to have a limited impact on vaccination inequities. By mid-December 2021, at the height of the winter Delta wave, and while the Omicron wave was looming, the French Prime Minister announced that the Sanitary Pass would become a Vaccine Pass, i.e. that a negative Covid-19 test would not provide a temporary QR code any longer for adults – making vaccination implicitly mandatory in France. The Vaccine Pass came into force on 24 January 2022.

This study aims to obtain further insights on the socio-economic, political and geographic factor associated with vaccination rates, and to evaluate the effect of the French domestic sanitary pass, by using nation-wide, exhaustive datasets.

## Methods

### Data

The French state health insurance service (Assurance Maladie) provides public datasets of vaccination rates in France. These datasets are based on aggregated individual data on beneficiaries of the national health insurance service who received health care in the past year. These exhaustive datasets are updated weekly, and are provided at the district scale nationally (EPCI: *Établissement public de coopération intercommunale*, an administrative level gathering multiple towns or cities) and at the suburban scale for the Paris, Lyon, and Marseille metropolitan areas. For this study, we focused on mainland France, because vaccination rates are much lower in oversea localities, and because determinants of vaccination rates are likely to differ in oversea localities compared to mainland ones. Our dataset included 1555 districts (1228 EPCIs and 327 districts at the suburban scale in Paris, Lyon, Marseille).

The vaccination dataset for mainland France encompasses about 64.5 million individuals (median district size 22310 inhabitants, interquartile range 11012–43038). The vaccination data are available by age class: 00–19, 20–39, 40–54, 55–64, 65–74, 75 and over. Population sizes for each locality and each age class are also provided.

We paired these vaccination data with three other datasets gathering socio-economic, political orientation and geographic variables.

Socio-economic data are provided by the French national statistics institute (INSEE), and are available at the same administrative levels as the vaccination data. We selected the most recent dataset available (year 2018). The different variables available in the dataset are classified by INSEE according to 8 categories (Activity, Education, Employment, Family, Housing, Immigration, Income, Population).

Latitude, longitude and surface data were extracted from open geographic datasets. We calculated from them four additional geographic indicators: distance to Paris, relative position along a South-East–North-West gradient, relative position along a South-West–North-East gradient, and local population density.

Political orientation data consisted of the results of the 2017 Presidential election in France, which we aggregated to reconstitute the same administrative levels as the vaccination dataset. This political dataset contains the proportions of votes for each of the 11 candidates of the first round, 2 candidates of the second round (Macron and Le Pen), and proportion of abstention at each round.

These three datasets comprised 312 indicators. We then removed those indicators with over 5% missing data, or with over 0.9 correlation with other indicators of the dataset, which left us with 141 indicators: 123 socio-economic indicators (Activity: n = 10; Education: n = 16; Employment: n = 25; Family: n = 20; Housing: n = 30; Immigration: n = 1; Income: n = 13; Population: n = 8); 6 geographic indicators; 12 political indicators.

### Analysis

Vaccination was accessible to all adults in France after 27 May 2021. It opened to teenagers (12-17 year olds) on 15 June 2021, and to children (5-11 year olds) on 22 December 2021. Because of this differential accessibility of vaccines, and because vaccine passport rules also differed for non-adults, we excluded the 00-19 age class from our analysis, and focused on vaccination rates among 20+ year-old individuals (hereafter “adults”).

For each indicator in our dataset, at each of the four chosen dates (weeks 2021-W27, 2021-W31, 2021-W35, 2021-W49, 2022-W03, 2022-W07), we considered the association between living in a district above the median of a that indicator and individual first-dose vaccination rates among adults. Odds ratios (OR) were computed from the output of a logistic regression.

To be able to compare predictors irrespective of the direction of the effect, we considered the maximum of `OR, 1/OR` (hereafter 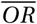). Note that vaccination data are at the individual level, and indicator data at the district level. The analysis is done at the individual level, with indicators characterizing the geographic districts in which individuals live.

For each date, we determined a significance threshold by computing odds ratios on 1000 random permutations of a predictor, and identifying the value of the 99% percentile odd ratios 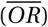 of these permuted data.

For representative indicators among the most statistically significantly associated ones, we estimated standardized vaccination rates among adults over time, for each decile of each indicator (treated as a factor). These estimations were obtained from a logistic model taking age class into account; adult vaccination rates were standarsized using an age distribution matching that of mainland France.

All analysis code is available at https://github.com/flodebarre/covid-passports-france (and will be stored on a permanent repository upon acceptance); analyses were done in R version 4.0.4 (2021-02-15).

## Results

We investigated the associations between each of the 141 indicators characterizing districts of residence, and the fact of having received at least one Covid-19 vaccine dose, on the whole population of mainland France. Two indicators were among the top five most associated one at all time points (see Figure 1): the share of local income coming from unemployment benefits (`Unemployment_Benef`; strongest association on 2022-01-23, *OR =* 0.716) and vote for the “anti-establishment” political party represented by the candidate Asselineau (`Asselineau`; *OR =* 0.712 on 2022-02-20). The three other most associated indicators did not change in the later dates that we considered, and were the proportion of immigrants in the district (`Immigrant`; *OR =* 0.713 on 2022-02-20), the district’s relative position along a North-West–South-East gradient (`NO-SE`; *OR =* 0.745 on 2021-12-12) and the proportion of overcrowded households (`Overcrowding_rate`; *OR =* 0.738 on 2022-01-23).

**Figure 1:**
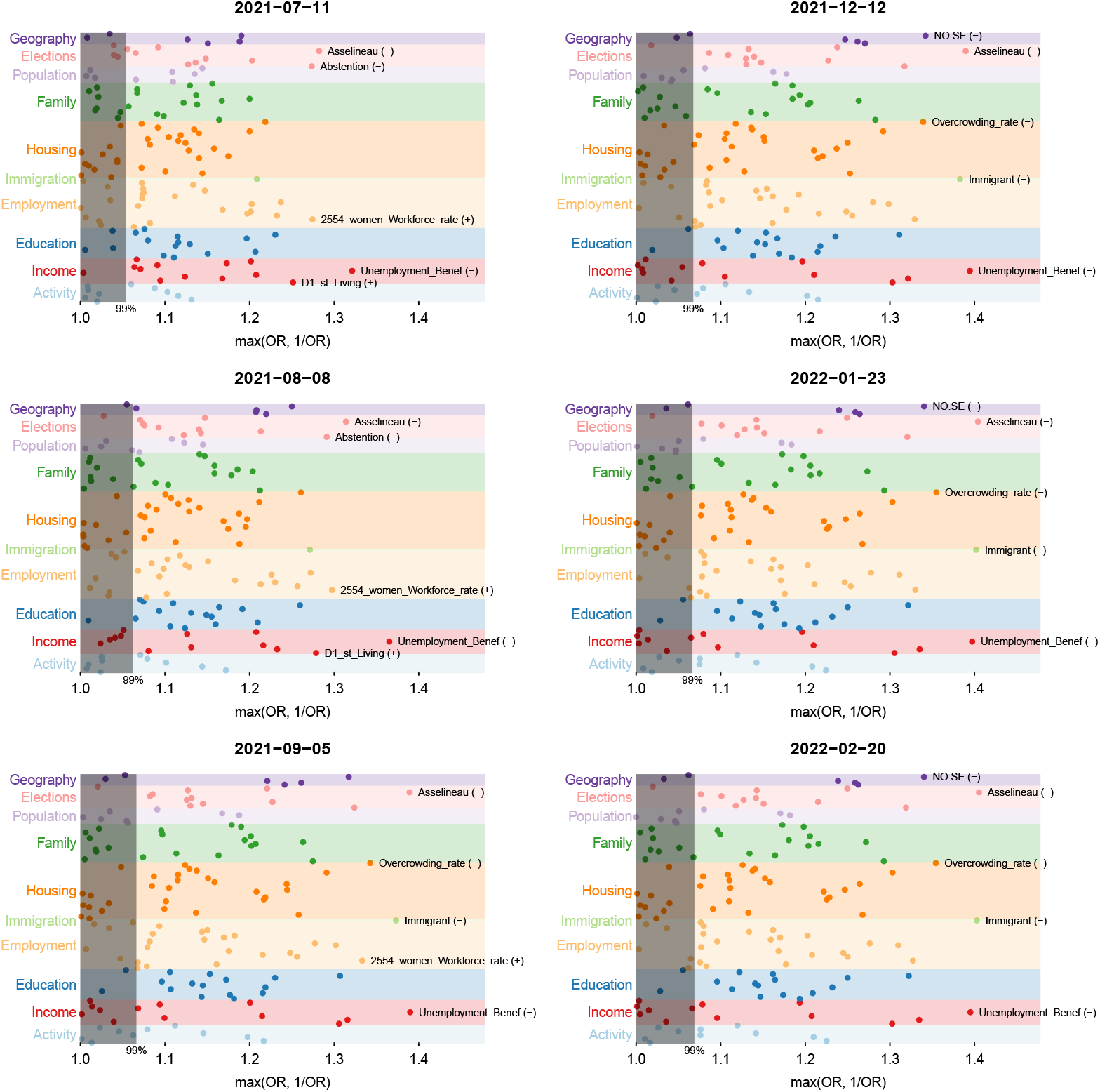
Manhattan plots of the Odds ratios for each of the indicator of our dataset, by date. Left column: around the Sanitary Pass implementation; right column: around the Vaccine Pass implementation. The top odds ratios are labelled at each time point; the symbol next to the name indicates the direction of the effect. The gray rectangle corresponds to the 99% percentile of odds ratios in the permuted data; points falling in the rectangle are considered as non-significant.

Our odds ratio calculations were based on a crude version of each indicator, which were dichotomized into values above or below the median of each indicator. To better visualize the effects (or lack thereof) of the sanitary and vaccine passes on vaccination rates over time, we computed age-adjusted vaccination rates over time, by decile of three of the most associated indicators, treated as factors (see Figure 2). The Sanitary Pass, implemented in the Summer 2021, led to an overall increase in vaccination rates; on the other hand, the Vaccine Pass, implemented in the end of 2021, did not affect the evolution of vaccination rates. Heterogeneities in vaccination rates persisted after both types of pass; vaccination rates gradually decrease by decile of each indicator, confirming the association of these indicators with vaccination rates without threshold effect. Of note, for the Unemployment and Asselineau vote indicators, the difference between the 9th and 10th deciles appears to be much larger than between the other consecutive deciles.

**Figure 2:**
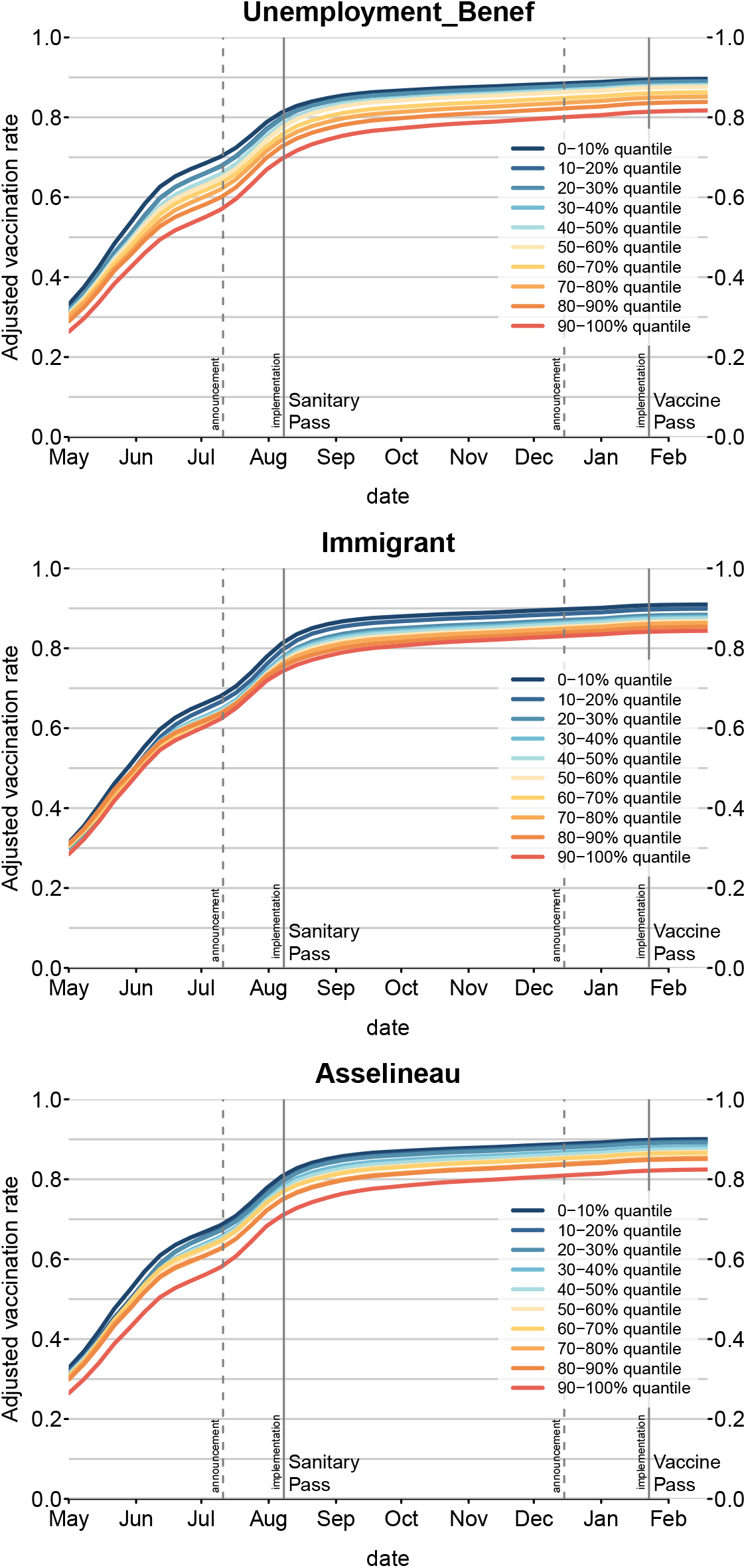
Age-adjusted vaccination rates among adults, over time, by decile of each indicator (presented by a color gradient). The vertical lines indicate the dates of announcements and implementations of the sanitary and vaccine passes.

Finally, historically under-vaccinated areas in France stand out as being less vaccinated against Covid-19, in particular the South-East region (see Figure 3).

**Figure 3:**
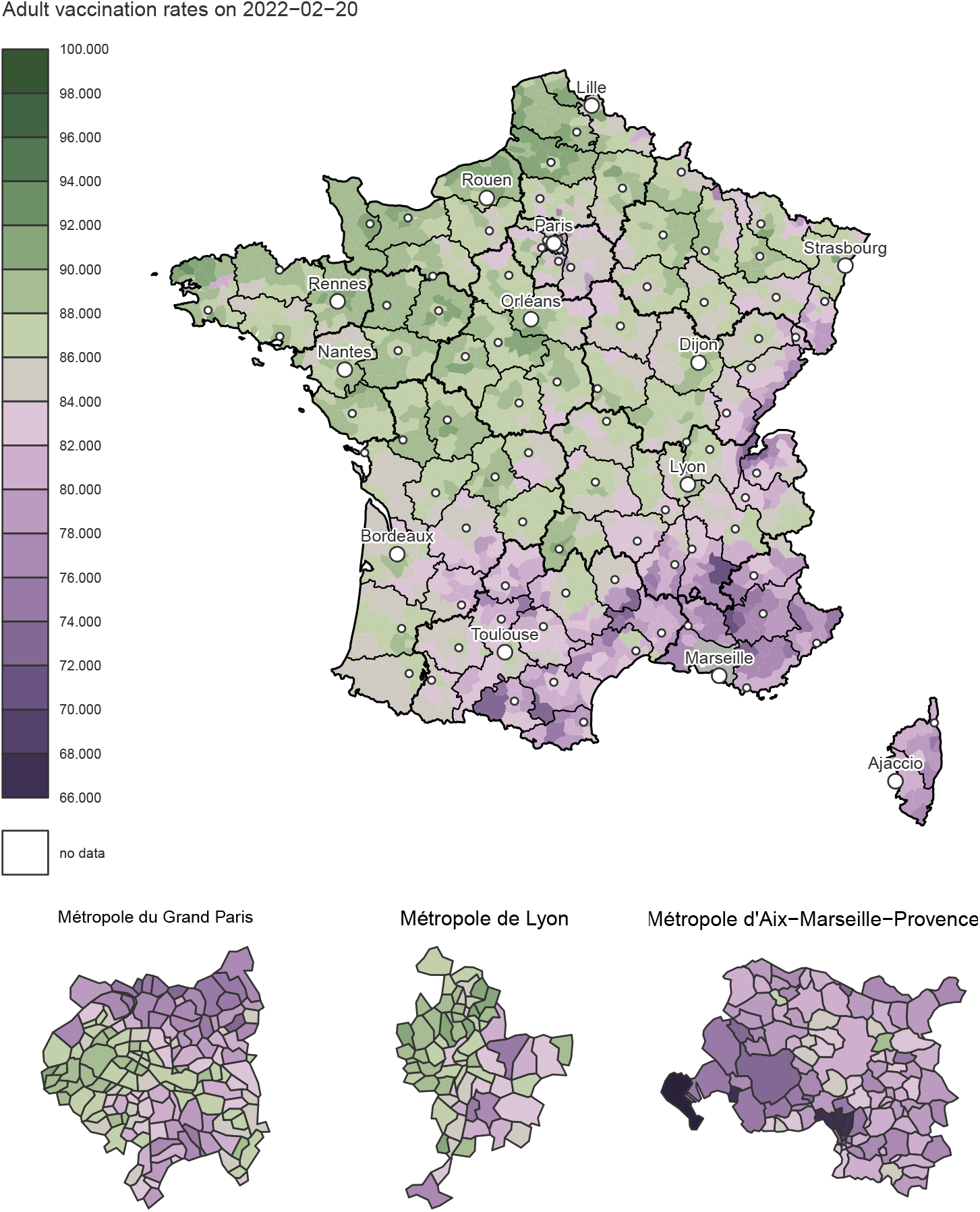
Adult vaccination rates by district of mainland France

## Discussion

Our results, based on exhaustive national datasets, indicate that the French sanitary pass, and the later vaccine pass, did not solve Covid-19 vaccination heterogeneities, but instead crystallized them. Indicators most associated with vaccination rates were associated to poverty, immigration, anti-establishment vote (or abstention), and a North-West – SouthEast contrast. For instance, the odds for an adult to still be unvaccinated by the end of February 2021 are about 1.4 times higher when living in the districts with higher than median value share of income coming from unemployment benefits, than when living in the districts with lower than median value.

The indicators associated to vaccination rates can be interpreted in the light of the dimensions of vaccine hesitancy (J. K. Ward et al. 2022). A first reason for vaccine hesitancy is complacency: not fully perceiving the benefit of vaccination or the risks of severe disease. While in this case a sanitary or vaccine pass may convince complacent individuals to get vaccinated, it is less efficient if the associated constrain is low. As the French domestic pass was associated to pay-for activities (restaurants, tourism), its persuading effect could be limited among poorer populations. This may explain the association of lower vaccination rate with poverty in the data that we analyzed: vaccination rates decrease as the share of local income coming from unemployment benefits (`Unemployment_Benef`) or the proportion of overcrowded households (`Overcrowding_rate`)) increase.

A second reason for vaccine hesitancy is confidence, i.e. trust in the vaccine, in the health care system, and more generally in the government (J. K. Ward et al. 2022; Lindholt et al. 2021). A survey conducted in July 2021 in France confirmed that trust in the government and trust in scientists were associated to higher odds to be vaccinated (Bajos et al. 2022). Votes for Mr Asselineau – which represented a minority of cast votes in 2017 in France (less than 1% overall) – can be interpreted as mistrust in the government (or more generally, against the establishment): This candidate for instance proposed that France exits the European Union, leave the Euro zone and reinstall the Franc currency; he was a proponent of hydroxychloroquin and ivermectin during the Covid-19 pandemic, and publicly expressed doubts about the safety of available Covid-19 vaccines. The association of higher proportion of votes for Mr Asselineau with lower vaccination rates can be interpreted as revealing a lack of confidence for the government. Noteworthily, among political indicators, the second strongest association is with abstention rates (higher abstention rates being associated to lower vaccination rates), again signaling higher distrust for institutions (J. K. Ward et al. 2020). Likewise, the lower vaccinations rates in the South-East of France can be interpreted as mistrust of the central government in Paris.

Finally, a third reason for vaccine hesitancy is convenience, that is, the availability and accessibility of the vaccines (J. K. Ward et al. 2022). During the first half of 2021, vaccination rate in France was mostly constrained by dose availability. Vaccination slots were to be booked online, and there was no general system for sending individual invitations to get vaccinated. It is therefore still possible that, in spite of some local outreach efforts, vaccine accessibility remained an issue, which may explain at least part of the association of lower vaccination rates with poverty. These accessibility issues may also explain the association we find between lower vaccination rates and living in a district with a high proportion of immigrants, which may for instance reveal language barriers. These associations of lower vaccination rates with more poverty and with higher proportions of immigrants in the district of residence are compatible with the results of a survey conducted in July 2021 in France (Bajos et al. 2022) on close to 81000 participants, which indicated that unvaccinated respondents were more likely to have lower income and more likely to belong to racialised minorities than vaccinated respondents

Relative position of the district of residency along a North-West–South-East gradient is also associated with vaccination probability, the South-East being less vaccinated. This geographic feature, already documented for other kinds of vaccination (Guimier 2021), have been shown to be the consequence of multiple determinants with a common consequence: a local climate of mistrust for the central Parisian power. Politically, anti-system votes (from the right as well as from the left) are traditionally concentrated in the South-East of France. Medically, General Practitioners (GPs) based in the South-East, and to a lesser extent those in the South-West, have been shown to tend to have a more negative opinion of vaccination than their colleagues practicing in the northern part of France (Gautier, Jestin, and Beck 2013). This greater skepticism influences GP practices and attitudes, resulting in a lesser degree of compliance with vaccination schedules than GPs in the northern half of France (Collange et al. 2015). Physical distance to the central government and institutions, based in Paris, coupled with a sense of belonging to a local community with a strong cultural identity, as is the case for example in the Marseille metropolis or in the Cévennes, play a role in indifference or mistrust towards institutions perceived as distant authorities (Guimier 2021). Finally, in and around the Marseille metropolis, the image of a rebellious territory was reinforced since the first months of the epidemic in France through the hypermediatized Pr Didier Raoult. Based in Marseille, he was a promoter of a controversial treatment against Covid-19 based on hydroxychloroquine and azithromycin (Schultz et al. 2022), and later held ambiguous positions regarding Covid-19 vaccination. He has become a local icon, thanks to his anti-system positions, and against the hostility of most of the medical world towards his work. All in all, around the city of Marseille, and more broadly in South-Eastern France, the climate of suspicion against Parisian institutions, which had long been rooted in the area, hardened during the Covid-19 crisis, and was associated with distrust of Covid-19 vaccines.

The design of our study offers several advantages. First, we used a data-driven approach, i.e. we did not focus on indicators that we *a priori* thought to be associated with vaccination rate. The indicators that we identified as the most associated with vaccination rates were not biased towards our previous knowledge or surveys about vaccine hesitancy. Secondly, the data that we used are real-world data on effective vaccination, and not vaccination intentions. Intentions to be vaccinated and realized vaccination may not always match, especially with the introduction of measures like the French Sanitary and Vaccine Passes. For instance, according to a survey conducted in the Fall 2021, the introduction of the sanitary pass led to an increase in the share of individuals reporting being “angry they had to be vaccinated” (J. K. Ward et al. 2022). From an immediate public health perspective, such as the limitation of the number of severe cases, realized vaccination rates are a more useful metric. Finally, the vaccination data we used are based on records of the national health insurance service: they cover 64.5 million individuals living in mainland France, and vaccination rates are not self-reported, which strongly limits reporting bias.

Still, the design of our study also presents limitations. While our vaccination data are at the individual level, the socio-economic, political and geographic indicators are at the district level, and must therefore be interpreted as such: for instance, we cannot not show that receiving unemployment benefits is associated with lower vaccination probability, but we find an association with lower vaccination probability and the fact of living in a district where a large share of income comes from unemployment benefits. In addition, although the different indicators are analysed independently in our study, their combinations may affect vaccination rates. For instance, the effect of mistrust in the government on vaccination refusal was shown to be even stronger among individuals from lower social classes than from higher social classes (Bajos et al. 2022). Finally, our data do not inform directly on the reasons for non-vaccination – e.g., whether it is hesitancy, refusal, or accessibility issues, which is why our approach is complementary to qualitative surveys.

To conclude, by emphasizing a differentiated use of COVID-19 vaccination according to a socio-economic gradient, our study confirms the strong impact of social inequalities on COVID-19. Previous research found that the most deprived areas have been disproportionately infected and hospitalized during the pandemic (Jannot et al. 2021; Bajos et al. 2021). We further show that poorer districts are also the least vaccinated and, hence, the most still at risk, despite the widely celebrated domestic sanitary pass. There is an urgent need to define new vaccination policies that truly address social inequities.

## Data Availability

This study involves only openly available data:
- INSEE: https://www.insee.fr/fr/statistiques/5359146#consulter
- Assurance Maladie: https://datavaccin-covid.ameli.fr/explore/dataset/donnees-devaccination-par-epci/https://datavaccin-covid.ameli.fr/explore/dataset/donnees-de-vaccination-parcommune/information/
- 2017 Presidential election: https://www.data.gouv.fr/fr/datasets/election-presidentielle-des-23-avril-et-7-mai-2017-resultats-definitifs-du-1er-tour-par-communes/#resource-d282e53a-d273-425d-95bb-8a0d7632c79a-header

https://datavaccin-covid.ameli.fr/explore/dataset/donnees-devaccination-par-epci/

https://datavaccin-covid.ameli.fr/explore/dataset/donnees-de-vaccination-parcommune/information/

https://www.data.gouv.fr/fr/datasets/election-presidentielle-des-23-avril-et-7-mai-2017-resultats-definitifs-du-1er-tour-par-communes/#resource-d282e53a-d273-425d-95bb-8a0d7632c79a-header

## Acknowledgements

We thank the producers of public datasets, in particular David Levy at INSEE and Antoine Rachas at Assurance Maladie.

## Contributions

ASJ and FD designed the study with inputs from all authors. EL extracted socio-economic and political orientation data at district scale and computed indicators. ASJ, EL and FD had full access to aggregated data used for this study and take responsibility for the integrity of the data. EL and FD did the analyses and takes responsibility for the accuracy of the data analysis. FD drafted the paper with the help of ASJ, MR. All authors critically revised the manuscript for important intellectual content and gave final approval for the version to be published.

## Data sources

- Assurance Maladie: https://datavaccin-covid.ameli.fr/explore/dataset/donnees-devaccination-par-epci/ https://datavaccin-covid.ameli.fr/explore/dataset/donnees-de-vaccination-parcommune/information/
- INSEE: https://www.insee.fr/fr/statistiques/5359146#consulter
- 2017 Presidential election: https://www.data.gouv.fr/fr/datasets/election-presidentielle-des-23-avril-et-7-mai-2017-resultats-definitifs-du-1er-tour-parcommunes/#resource-d282e53a-d273-425d-95bb-8a0d7632c79a-header

